# Genome-wide reduction in chromatin accessibility and unique transcription factor footprints in endothelial cells and fibroblasts in scleroderma skin

**DOI:** 10.1101/2020.06.24.20138040

**Authors:** Pei-Suen Tsou, Pamela J. Palisoc, Mustafa Ali, Dinesh Khanna, Amr H Sawalha

## Abstract

Systemic sclerosis (SSc) is a rare autoimmune disease of unknown etiology characterized by widespread fibrosis and vascular complications. We utilized an assay for genome-wide chromatin accessibility to examine the chromatin landscape and transcription factor footprints in both endothelial cells (ECs) and fibroblasts isolated from healthy controls and patients with diffuse cutaneous (dc) SSc. In both cell types, chromatin accessibility was significantly reduced in SSc patients compared to healthy controls. Genes annotated from differentially accessible chromatin regions were enriched in pathways and gene ontologies involved in the nervous system. In addition, our data revealed that chromatin binding of transcription factors SNAI2, ETV2, and ELF1 was significantly increased in dcSSc ECs, while recruitment of RUNX1 and RUNX2 was enriched in dcSSc fibroblasts. Significant elevation of *SNAI2* and *ETV2* levels in dcSSc ECs, and *RUNX2* levels in dcSSc fibroblasts were confirmed. Further analysis of publicly available ETV2-target genes suggests that ETV2 may play a critical role in EC dysfunction in dcSSc. Our data, for the first time, uncovered the chromatin blueprint of dcSSc ECs and fibroblasts, and suggested that neural-related characteristics of SSc ECs and fibroblasts could be a culprit for dysregulated angiogenesis and enhanced fibrosis. Targeting these pathways and the key transcription factors identified might present novel therapeutic approaches for this disease.

## Introduction

Systemic sclerosis (scleroderma, SSc) is a chronic debilitating disease characterized by immune activation, vascular injury, and enhanced accumulation of extracellular matrix proteins in many organs. The associated high mortality and morbidity for this disease is due to lack of effective treatment options to modify disease progression, with the exception of autologous hematopoietic stem cell transplant, which is restricted to a small proportion of patients and can be associated with significant adverse events.

Although the etiology of SSc is poorly understood, work from our group and others have generated evidence that strongly implicate epigenetic dysregulation in the pathogenesis of SSc^1^. This is supported by the low concordance rate for SSc in twins compared to other autoimmune diseases^2^. Indeed, we have shown that in both endothelial cells (ECs) and fibroblasts isolated from SSc skin, epigenetic mechanisms are critically involved in their disease phenotype. Distinct differences in genome-wide DNA methylation pattern were reported in fibroblasts isolated from limited cutaneous and diffuse cutaneous (dc) SSc patients^3^. The overexpressed methyl-CpG-binding protein 2 (MeCP2) in SSc fibroblasts appeared to be anti-fibrotic, in part mediated by its effect on target genes including *PLAU, NID2*, and *ADA*^4^. In addition to DNA methylation, changes in histone-modifying enzymes, such as the histone methyltransferase enhancer of zeste homolog 2 (EZH2), has also been implicated in promoting fibrosis in SSc fibroblasts^5^. We also showed that histone deacetylase 5 (HDAC5) and EZH2 were both upregulated in SSc ECs and posed detrimental effects on angiogenesis in these cells^5,6^.

In this study we provide a comprehensive analysis of chromatin accessibility patterns in SSc utilizing an assay for transposase-accessible chromatin with sequencing (ATAC-seq) with high-read density, allowing for a detailed analysis of transcription factor recruitment across the genome in skin-derived SSc ECs and fibroblasts compared to healthy controls.

## Methods

### Scleroderma patients and controls

Both male and female patients diagnosed with dcSSc were included in this study. All patients recruited for this study met the 2013 ACR/EULAR criteria for the classification of SSc^7^. Age-, sex-, and ethnicity-matched healthy controls were recruited (age 55.0 ± 3.2 years, mean ± SEM; 4 males and 19 females). All patients had dcSSc, including 5 males and 19 females. The age of the patients with SSc was 58.3 ± 3.0 years (mean ± SEM), and the disease duration was 3.0 ± 0.6 years (mean ± SEM). Their skin scores ranged from 0 to 37 with a mean of 20.0 ± 2.1 (mean ± SEM). All patients had Raynaud’s phenomenon, 5 had active digital ulcer, and 20 had pulmonary involvement at the time of biopsy. The detailed information of the patients and controls included in this study is shown in **Supplemental Table 1**. Subjects were recruited from the University of Michigan Scleroderma Program. The institutional review board approved this study and all participants signed an informed consent prior to enrollment.

**Table 1:** Gene ontology analysis of genes associated with differential chromatin accessibility in SSc ECs compared to the normal ECs. Only the top 10 ontology terms in each category are depicted.

| Gene Ontology Term | Number of Genes involved | Adjusted P Value |
| --- | --- | --- |
| **<u>Biological Process:</u>** |  |  |
| Cell projection organization | 66 | 1.30E-15 |
| Neuron differentiation | 53 | 1.57E-10 |
| Neurogenesis | 57 | 7.23E-10 |
| Neuron development | 42 | 1.02E-07 |
| Cell projection assembly | 28 | 4.37E-07 |
| Positive regulation of cellular biosynthetic process | 57 | 9.02E-07 |
| Axon development | 25 | 5.47E-06 |
| Locomotion | 54 | 1.07E-05 |
| Cellular macromolecule localization | 53 | 1.07E-05 |
| Positive regulation of nucleobase-containing compound metabolic process | 51 | 2.32E-05 |
| **<u>Cellular Component:</u>** |  |  |
| Synapse | 46 | 1.14E-07 |
| Chromosome | 51 | 1.23E-06 |
| Microtubule cytoskeleton | 39 | 8.59E-06 |
| Neuron projection | 40 | 1.57E-05 |
| Nuclear chromosome | 39 | 1.57E-05 |
| Post-synapse | 25 | 2.42E-05 |
| Nucleus | 31 | 2.77E-05 |
| Chromatin | 37 | 2.77E-05 |
| Transferase complex | 28 | 2.77E-05 |
| Microtubule organizing center | 27 | 5.13E-05 |
| **<u>Molecular Function:</u>** |  |  |
| Transcription regulator activity | 52 | 7.67E-07 |
| Ribonucleotide binding | 55 | 7.67E-07 |
| Adenyl nucleotide binding | 47 | 2.13E-06 |
| Drug binding | 48 | 1.48E-05 |
| Kinase activity | 29 | 1.48E-05 |
| Protein kinase activity | 25 | 1.52E-05 |
| Protein dimerization activity | 40 | 1.52E-05 |
| Calcium ion binding | 27 | 2.25E-05 |
| Signaling receptor binding | 44 | 4.69E-05 |
| Transferase activity transferring phosphorus containing groups | 31 | 4.69E-05 |

### Cell culture

Two 4 mm punch biopsies from the distal forearm of healthy volunteers and SSc patients were obtained for cell isolation. Skin digestion and cell purification were described previously with slight modification^5, 8^. After skin digestion, cells were grown to 75% confluency before purification. ECs were purified through positive selection using the CD31 MicroBead Kit (Miltenyi Biotech) while negatively selected cells eluted were fibroblasts. A portion of the collected fibroblasts (passage 0) were lysed and underwent tagmentation as mentioned below. The remainder fibroblasts were cultured in RPMI supplemented with 10% fetal bovine serum (FBS) and antibiotics.

Fibroblasts between passages 3 and 6 were used in subsequent studies. ECs were maintained in EBM-2 media supplemented with EGM-2 MV Microvascular Endothelial SingleQuots Kit from Lonza. These cells underwent several rounds of purification during expansion and cells with passages between 2 and 6 were used for ATAC-seq library preparation as well as subsequent experiments in this study. Since the EBM-2/EGM-2 media contains multiple growth factors such as VEGF and FGF2, as well as antioxidant vitamin C, to steer ECs to a less activated state, cells were cultured in EBM media supplemented with EGM-MV Microvascular Endothelial Cell Growth Medium BulletKit supplements (containing bovine brain extract, Lonza) for at least 24 hours before ECs were collected for library preparation or experiments.

### Assay for transposase-accessible chromatin using sequencing

ATAC-seq was performed to assess genome-wide chromatin accessibility as previously described^6, 9^. For both fibroblasts and ECs (from 6 patients and 6 controls), 50,000 cells were used for the Tn5 transposition reaction, and DNA libraries were prepared. Libraries that passed the quality control were sequenced with 50bp, pair-end reads on the Illumina NovaSeq 6000 platform at a read depth of approximately 150 million reads/sample. Reads were pre-processed to remove adapter reads, and then aligned to the reference genome. Chromatin accessibility peaks were identified and then differential peaks between healthy controls and dcSSc were identified using edgeR R Bioconductor package^10^ (v3.26.5). The Benjamini–Hochberg method was used to adjust p-values from multiple testing and only genes with adjusted p-values < 0.05 and |logFoldChange| ≥ 1 were used for subsequent analysis. We used the HINT tool in the Regulatory Genomics Toolbox (rgt-hint) to perform footprinting and motif binding analysis^11^. Briefly, for each condition, the reads among the replicates are pooled and tag counts are determined within the merged peaks. Next, motifs from the JASPAR database^12^ were queried against the footprints found within each condition. Finally, a differential score-the activity score, representing the transcription factor binding activity and the openness of the surrounding chromatin is calculated and visualized between pairs of conditions (normal-dcSSc). In this case, positive activity scores represent more binding of transcription factors in normal cells, and negative scores represent more binding of transcription factors in dcSSc cells.

### Bioinformatics analysis

Functional enrichment analyses for annotated genes were performed using Molecular Signatures Database (v7.1)^13, 14, 15^ and ToppGene^16^. The canonical pathways were derived using gene sets from pathway databases, including BioCarta, KEGG, PID, and Reactome. To analyze potential transcription factors regulating a gene set, iRegulon (v1.3) was used^17^. Further functional enrichment analyses to generate networks for visualization were performed using ClueGO (v2.5.7)/CluePedia (v1.5.7)^18, 19^ and Cytoscape (v3.8.0)^20^. The position weight matrices for each transcription factor were acquired from the JASPAR database (2020 release)^12^.

### mRNA extraction and qRT-PCR

The Direct-zol™ RNA MiniPrep Kit (Zymo Research) was used to extract RNA before it was converted to cDNA using the Verso cDNA synthesis kit (Thermo Scientific). Primers for human *RUNX1, RUNX2, ETV2, ELF1, SNAI1, SNAI2*, or β*-actin* were mixed with Power SYBR Green PCR master mix (Applied Biosystems), and the cDNA was quantified using the ViiA™ 7 Real-Time PCR System.

### Statistics analysis

Results were expressed as mean ± S.D except stated otherwise. To determine the differences between the groups, Mann–Whitney U test was performed using GraphPad Prism version 8 (GraphPad Software, Inc). *P*-values of less than 0.05 were considered statistically significant.

## Results

### Chromatin accessibility is broadly decreased in dcSSc cells

The ATAC-seq libraries were sequenced to an average of 150 million reads per sample. All ATAC-seq libraries yielded fragment lengths with the expected fragment distribution and clear nucleosome phasing (**Figure 1A**). The accessible regions identified were mostly enriched within 1 kb of the TSS (**Figure 1B**), which is consistent with presence of regulatory elements in these regions. We next compared our ATAC-seq data with ENCODE open chromatin data generated using DNase-seq, as well as ChIP-seq data targeting H3K27ac, which marks active enhancers (**Figure 1C**). There were no publicly accessible data for H3K27ac marks in human dermal ECs therefore we also included tracks generated from human umbilical vascular ECs (HUVECs). The ATAC-seq peaks from both SSc ECs and fibroblasts overlapped with ENCODE peaks, further supporting the quality of our data.

**Figure 1:**
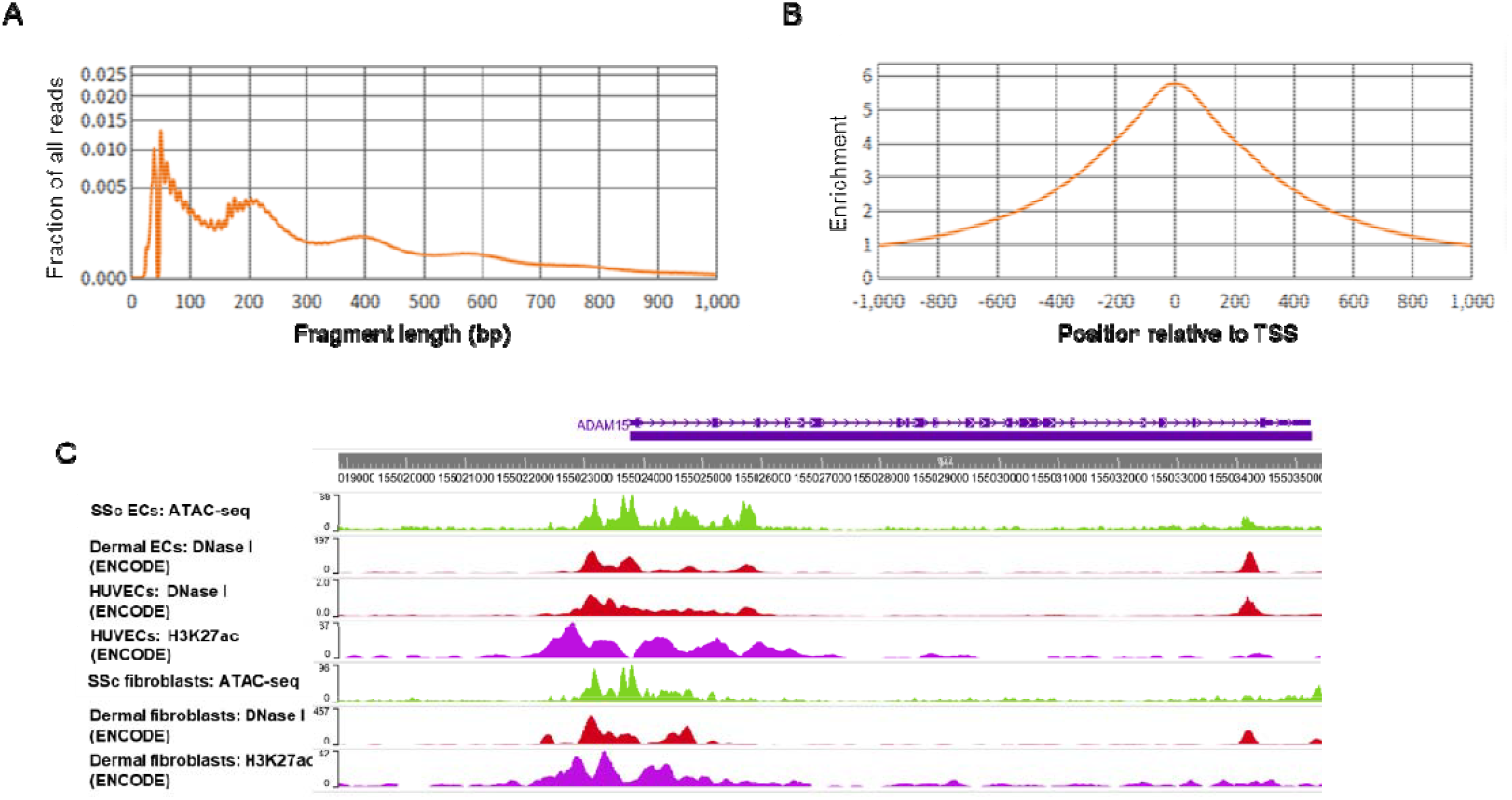
Overview of the ATAC-seq results in dermal ECs and fibroblasts. (**A**) Fragment size distributions of ATAC-seq data from a representative EC sample showing clear nucleosome phasing. The first peak represents the open chromatin, subsequent peaks represent mono-, di- and tri-nucleosomal regions. (**B**) Average plot of a representative EC sample showing enrichment of ATAC-seq signals around transcription start sites (TSS). (**C**) Representative sequencing tracks for the *ADAM15* locus show distinct ATAC-seq peaks at the promoter and the known enhancer in SSc ECs and fibroblasts. The ATAC-seq peaks identified overlapped with ENCODE open chromatin data generated using DNase-seq in human dermal ECs, fibroblasts, or human umbilical vascular ECs (HUVECs). ATAC-seq peaks also overlapped with ENCODE H3K27ac peaks marking active enhancers in HUVECs and dermal fibroblasts.

Chromatin accessibility in SSc ECs and fibroblasts was overall lower than that in normal ECs and fibroblasts, potentially reflecting the disease state of SSc cells (**Figure 2 and 3**). ATAC-seq identified a total of 558 regions that are differentially accessible in SSc ECs compared to normal ECs, among them 97 regions showed increased chromatin accessibility while 461 regions had decreased chromatin accessibility in SSc cells (**Figure 2A**). This corresponds to 74 and 390 genes in these regions, respectively. **Figure 2B** depicts the genome tracks of the *CDH2* and *NLGN1* loci as two examples showing the regions that were differentially less accessible in dcSSc ECs compared to normal ECs, as indicated by the decrease in peak intensity. To examine the distribution of the differentially accessible chromatin regions, we identified enriched regions and mapped them to different genomic features. We found that the genomic location of the differentially accessible regions in ECs were enriched in introns (**Figure 2C**). Less accessible peaks in SSc ECs were enriched in the promotor regions. As shown in **Figure 2C**, among the putative more accessible regions (opening), 11% were located in proximal promoters (within 1 kb of the transcription start site, TSS), 11% in distal promoters (1 to 5 kb of TSS), 24% in exons, 41% in introns, 8% in 5’-UTR, and 5% in 3’-UTR. In the less accessible regions (closing), 21% were located in proximal promoters, 9% in distal promoters, 21% in exons, 28% in introns, 20% in 5’-UTR, and 1% in 3’-UTR.

**Figure 2:**
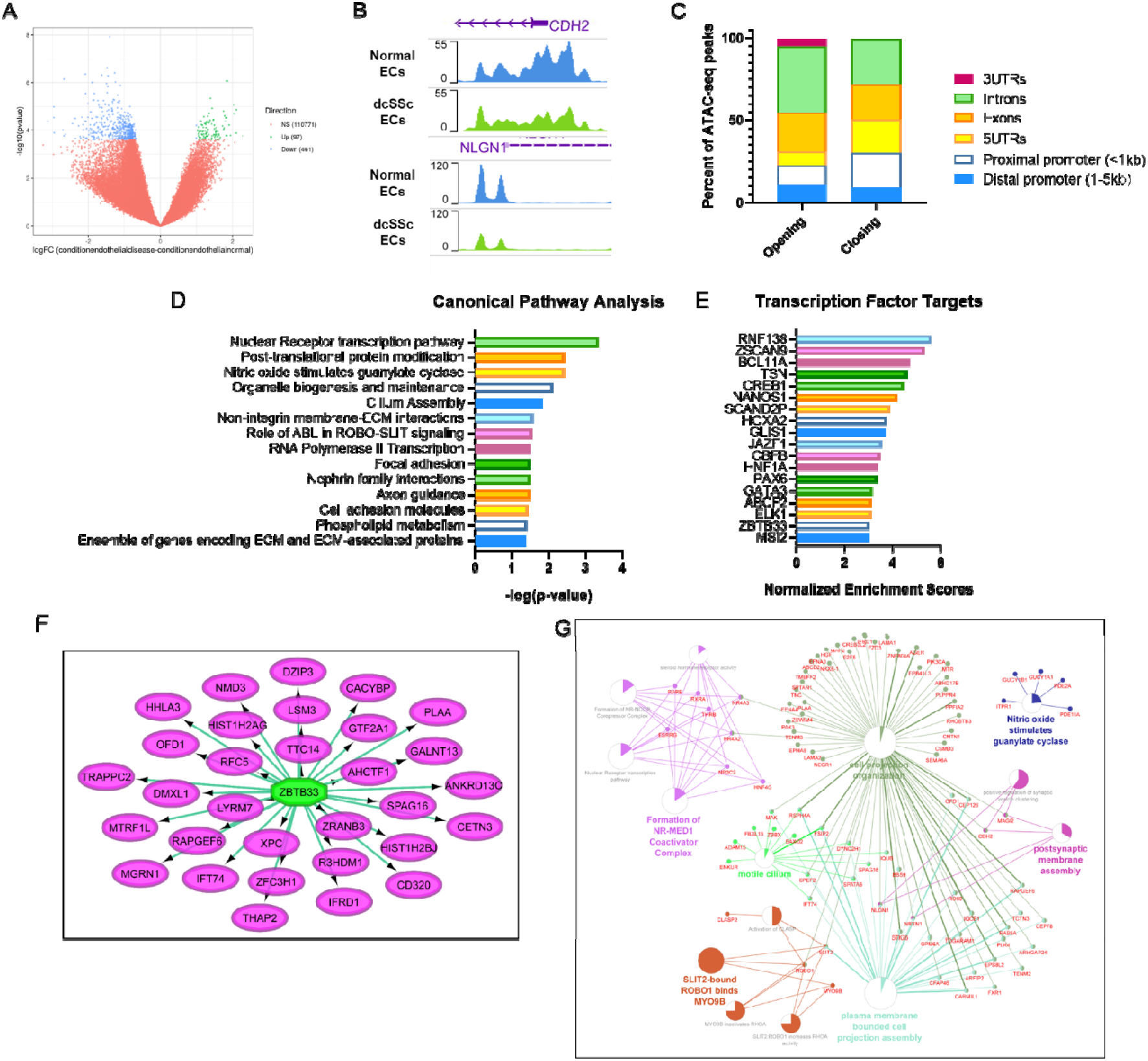
ATAC-seq captures chromatin accessibility in dcSSc ECs. (**A**) Volcano plot of differentially accessible genes between normal and SSc ECs. Values with logFC > 1 or logFC < −1 and adjusted P-value < 0.05 are highlighted in green and blue, respectively. (**B**) Representative ATAC-seq tracks at *CDH2* and *NLGN1* gene loci in normal and dcSSc ECs. Both genes are neuronal-related genes. (**C**) Genomic distributions of differentially accessible chromatin regions identified by ATAC-seq in ECs. Each color represents a genomic feature, as shown in the right part of the plot. Opening and closing refers to higher and lower chromatin accessibility in dcSSc relative to normal ECs, respectively (**D**) Pathway enrichment analysis of loci annotated to all differentially accessible regions in dcSSc ECs. (**E**) Gene regulatory network analysis for transcription factor targets using iRegulon, showing the top 18 most significant enrichments. Normalized Enrichment Score corresponds to a false discovery rate between 3% and 9%. (**F**) An example of a gene regulatory network showing differentially accessible genes identified by ATAC-seq in dcSSc ECs that are targets of the transcription factor ZBTB33. (**G**) Network analysis representing genes annotated to differentially accessible chromatin regions in dcSSc ECs compared to controls. The node size corresponds with the significance of the term it represents within the network. Colored portions of the pie charts represent the number of genes identified from the ATAC-seq experiment that are associated with the term.

**Figure 3:**
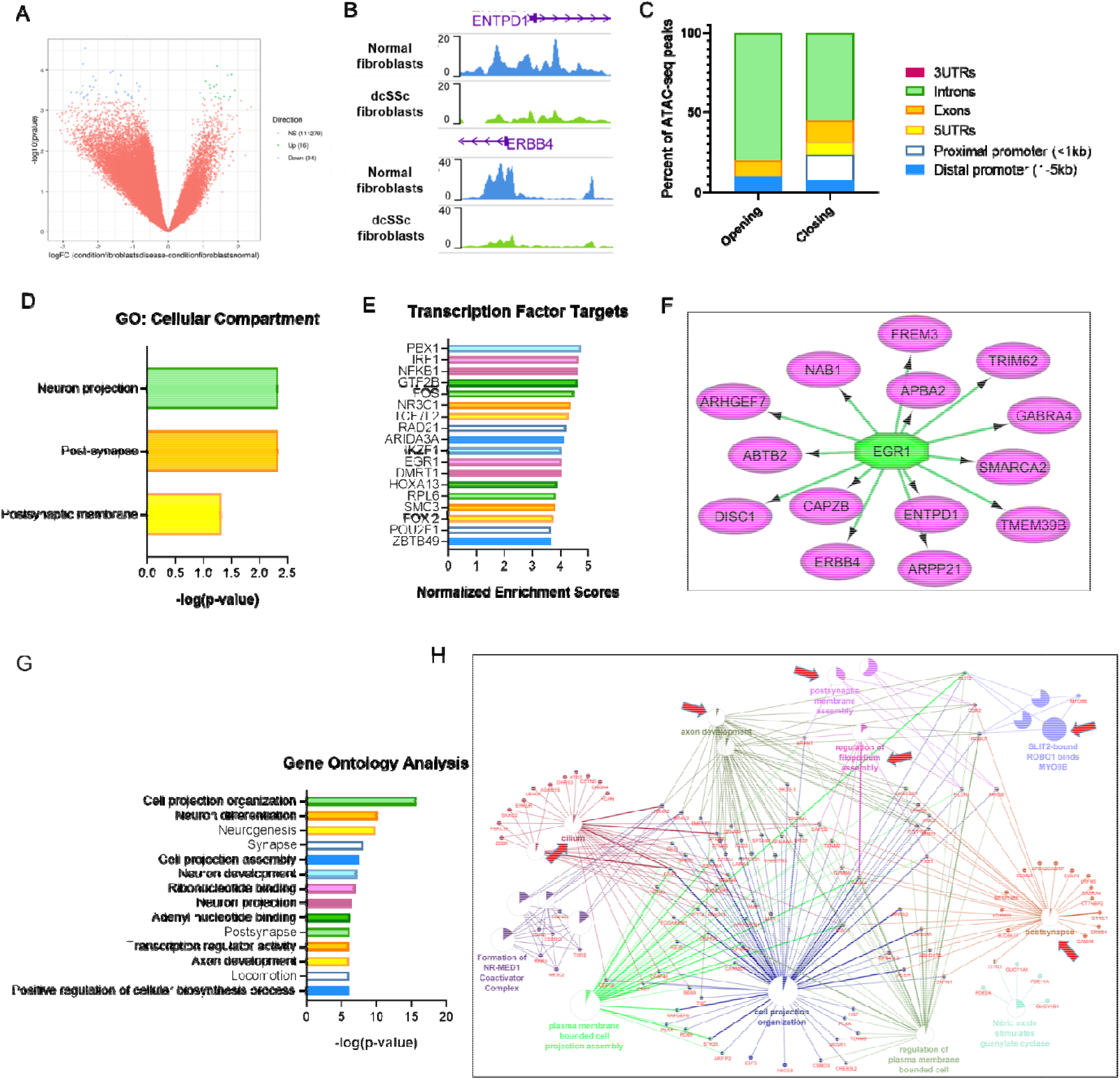
Analysis of chromatin accessibility profiles in dcSSc fibroblasts. (**A**) Volcano plot of differentially accessible genes between normal and SSc fibroblasts. Values with logFC > 1 or logFC < −1 and adjusted P-value < 0.05 are highlighted in green and blue, respectively. (**B**) Representative ATAC-seq tracks at *ENTPD1* and *ERBB4* loci in normal and dcSSc fibroblasts. (**C**) Genomic distributions of differentially accessible chromatin regions identified by ATAC-seq in fibroblasts. Each color represents a genomic feature, as shown in the right part of the plot. Opening and closing refer to higher and lower chromatin accessibility in dcSSc relative to normal fibroblasts, respectively (**D**) Gene ontology analysis of loci associated with differentially accessible regions in dcSSc fibroblasts. (**E**) Gene regulatory network analysis for transcription factor targets using iRegulon, showing the top 18 most significant enrichments. Normalized Enrichment Score corresponds to a false discovery rate between 3% and 9%. (**F**) An example of a gene regulatory network showing differentially accessible genes identified by ATAC-seq in dcSSc fibroblasts that are targets of the transcription factor EGR1. (**G**) Major gene ontology category annotations of genes associated with differentially accessible chromatin in both SSc ECs and fibroblasts. (**H**) Network analysis of differentially accessible chromatin regions identified in both dcSSc ECs and fibroblasts. The node size corresponds with the significance of the term it represents within the network. Colored portions of the pie charts represent the number of genes identified from the ATAC-seq experiment that are associated with the term. The clusters related to the nervous system are highlighted by arrows.

To gain insight into the functions of the genes that are located in differentially accessible regions in SSc ECs, we performed functional enrichment analyses using Molecular Signatures Database and found that the top 3 pathways that these genes were significantly associated with included nuclear receptor transcription, post-translational protein modification, and NO-guanylate cyclase (**Figure 2D** and **Supplemental Table 2**). Gene ontology analysis revealed that these genes were significantly associated with cell projection organization, chromosome, transcription regulator activity, and many neuronal-related GO terms (**Table 1**). We also explored genes representing potential targets of regulation by transcription factors, with several transcription factors identified including RNF138, ZSCAN9, and BCL11A, to name a few (**Figure 2E**). An example of gene regulatory network is shown in **Figure 2F**. 31 gene annotated from differential chromatin accessible regions from dcSSc ECs were targets of transcription factor ZBTB33, which is a transcription factor also identified in the HINT-ATAC analysis in ECs shown below. To further capture the relationships among the pathways and gene ontology terms, we used ClueGO/CluePedia to generate a network plot comprising a subset of enriched terms with the best p-values (**Figure 2G**), showing the gene clusters of enriched pathways for nuclear receptors, NO-cGMP, cilium, the nervous system, and cellular compartment organization.

In fibroblasts, we identified 50 genetic loci with differential chromatin accessibility between normal and dcSSc fibroblasts. 16 and 34 loci demonstrated increased and decreased chromatin accessibility, respectively, in dcSSc compared to normal fibroblasts (**Figure 3A**). This corresponds to 9 and 24 genes in these regions. These data are consistent with overall reduced chromatin accessibility in the cells from dcSSc patients. The genome tracks of two selected genes, *ENTPD1* and *ERBB4*, were selected to show the decrease in chromatin accessibility in dcSSc fibroblasts when compared to normal fibroblasts (**Figure 3B**). Similar to the distribution of chromatin regions in SSc ECs, the majority of peaks were detected in the introns in fibroblasts (**Figure 3C**). In the more accessible regions (opening), 10% were located in distal promoters, 10% in exons, 80% in introns, while none in proximal promoter, 5’-UTR, and 3’-UTR. In the less accessible regions (closing), 16% were located in proximal promoters, 8% in distal promoters, 13% in exons, 55% in introns, 8% in 5’-UTR, and 0% to 3UTR. The 33 genes that were located in differential chromatin accessibility regions were subjected to gene ontology analysis and they were enriched in neuron projection, post-synapse, and postsynaptic membrane (**Figure 3D** and **Supplemental Table 3**).

They were also potential targets of transcription factors PBX1, IRF1 and NFKB1, among others (**Figure 3E**). A gene regulatory network was presented as **Figure 3F**; 14 genes from the ATAC-seq data set from dcSSc fibroblasts were targets of transcription factor EGR1, which is an enriched transcription factor in fibroblasts independently identified from our HINT-ATAC analysis shown below.

From our analysis above, the genes located in differentially accessible regions from both ECs and fibroblasts appeared to be enriched in pathways that are involved in the nervous system. Indeed, gene ontology analysis of loci associated with differentially accessible regions from both cell types revealed a significant enrichment in neuron differentiation, neurogenesis, synapse, neuron development, neuron projection, post-synapse, and axon development; 8 out of the top 14 most significant gene ontologies (**Figure 3G**). The gene network also shows significant clustering of gene ontologies related to the nervous system, as indicated by the arrows (**Figure 3H**).

### Transcription factor binding analysis at open chromatin regions in SSc cells

Within the accessible chromatin regions, putative transcription factors can be detected since the narrow genomic regions occupied by transcription factors are protected from Tn5. We used HINT-ATAC to identify enriched putative transcription factor motifs in the open chromatin regions of both ECs and fibroblasts comparing healthy cells to SSc cells. In ECs, TFDP1, ZBTB33, E2F4, NFYA, and HINF, among others, bind more in normal ECs compared to SSc ECs, as indicated by their positive activity score (**Figure 4A** and **Supplemental Table 4**). In dcSSc ECs, we observed significantly more binding of motifs of ETV2, SNAI2, and ELF1 compared to healthy ECs (**Figure 4A** and **Supplemental Table 4**). The footprints of these transcription factors are shown in **Figure 4B**. The putative transcription factors differentially recruited in ECs between dsSSc patients and healthy controls, identified in the HINT-ATAC analysis, were enriched in pathways critical in telomere maintenance, TGFβ-regulated pathways, nerve growth factor (NGF) pathways, and P53 pathways (**Figure 4C** and **Supplemental Table 5**).

**Figure 4:**
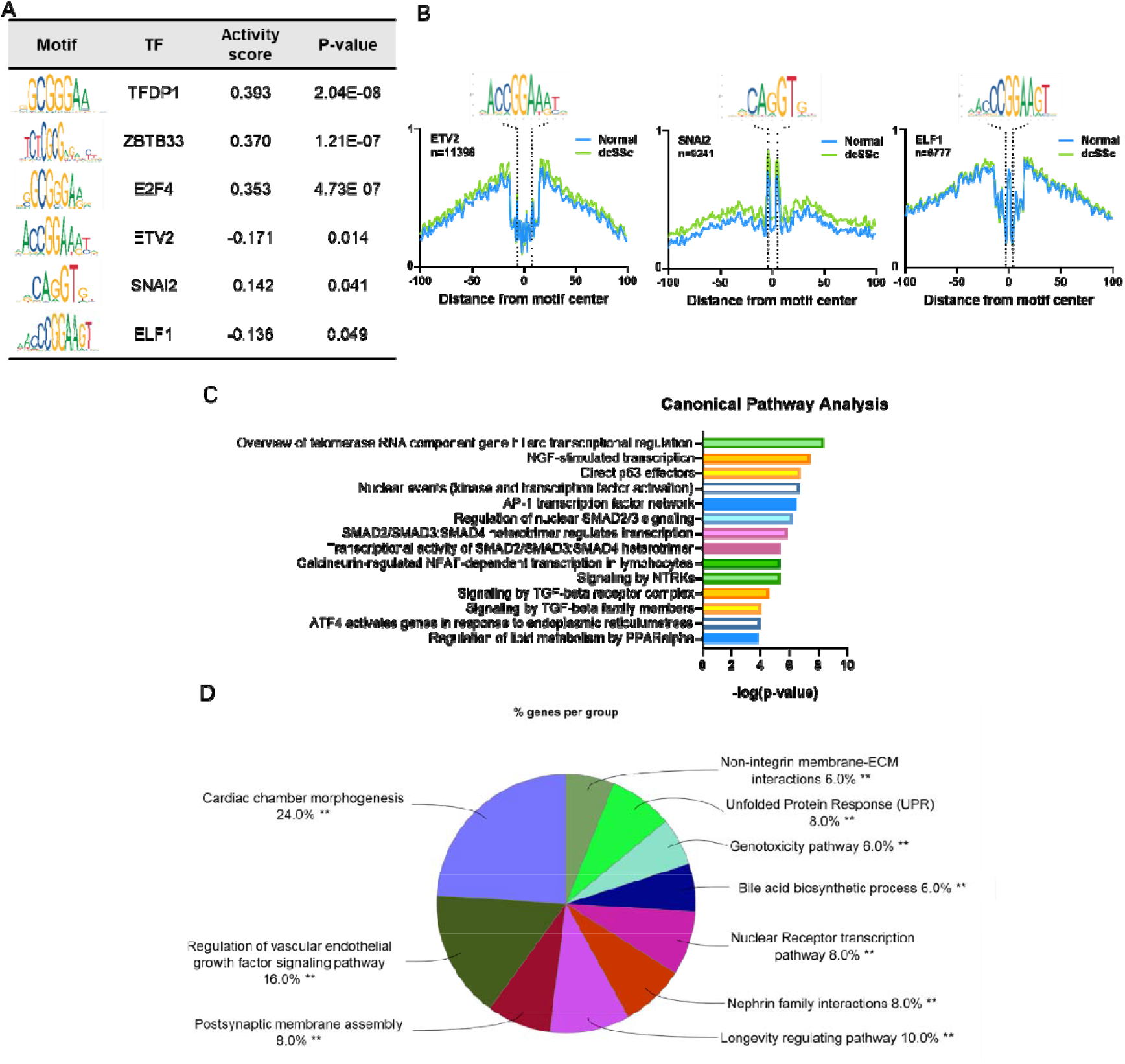
The most enriched transcription factor motifs and their target genes in SSc ECs. (**A**) HINT-ATAC motif analysis of regions in normal and dcSSc ECs. The top 3 transcription factors with significant differential activity values in ECs are shown. The activity score indicates the difference in activity in normal compared to dcSSc ECs (Normal-dcSSc). A positive score indicates increased transcription factor binding in normal ECs, and a negative score indicates increased binding in dcSSc ECs. (**B**) The footprint profiles of ETV2, SNAI2, and ELF1 generated from ECs. (**C**) Pathway analysis of the putative transcription factors identified in ECs. The top 14 are shown. (**D**) Pathway analysis of differentially accessible genes in dcSSc ECs that overlap with ETV2-target genes reported in Liu et al.

To help us understand the regulatory role of the transcription factors enriched in dcSSc ECs, we extracted gene sets from the TRANSFAC Predicted Transcription Factor Targets and CHEA Transcription Factor Targets databases^21, 22^ for SNAI2 and ELF1, respectively. For ETV2, the gene list generated via ChIP-seq in *in vitro* differentiated embryonic stem cells was used^23^. Pathway analysis revealed that SNAI2-target genes are enriched in nuclear receptor transcription and SUMOylation, and that ETV2-target genes are enriched in nervous system development and axon guidance (**Supplemental Table 6**). We also examined the degree of overlap between differentially accessible genes in ECs between patients and controls and the identified target genes for SNAI2, ETV2, or ELF1. Between the SNAI2-target genes and the differentially accessible ATAC-seq-annotated genes identified, there were 4 genes that overlapped (*CACYBP, RXRA, SP4*, and *THRB*). Between the ELF1-target gene set and the differentially accessible genes there were 17 overlapping genes (*ARRDC2, AZI2, CCNG1, CD320, EAF2, EDRF1, ESPL1, FAM171A2, GHITM, KCNN1, MYO9B, PDE4A, RAPGEF6, SLC12A4, SPTY2D1, TTC14, and XPC*). For ETV2, there were 89 genes overlapping between genes identified by ATAC-seq and the published ChIP-seq data (**Supplemental Figure 1**). These include genes involved in neural-related pathways such as *CDH2, NLGN1, ROBO1*, and *SLIT2*, as well as genes involved in longevity regulating pathway and vascular endothelial growth factor pathway (**Figure 4D**). Based on these results, the significant enrichment of genes involved in the nervous system identified in the differential chromatin accessibility analysis between SSc and normal ECs (**Figure 2D** and **2G**) might be driven by enriched ETV2 binding in dcSSc ECs.

In normal fibroblasts, HINT-ATAC analysis identified more binding of NRF1, TCFL5, ZBTB33, E2F4, and TFDP1, among other transcription factors, compared to fibroblasts from dcSSc patients (**Figure 5A** and **Supplemental Table 7**). In dcSSc fibroblasts, significant enrichment of RUNX1 and RUNX2 was observed compared to normal fibroblasts (**Figure 5A** and **Supplemental Table 7**). The footprints are shown in **Figure 5B**. When we performed pathway enrichment analysis, the transcription factors with differential recruitment across the genome between dcSSc and normal fibroblasts were significantly enriched in TGFβ-regulated pathways, NGF pathways, signaling by neurotrophin receptors (NTRKs), and telomere maintenance (**Figure 5C** and **Supplemental Table 8**). We extracted gene sets from the CHEA Transcription Factor Targets database^21^ for RUNX1 and RUNX2. There were 1263 overlapping genes between the two gene sets. Pathway analysis examining RUNX1 and RUNX2-target genes revealed enriched pathways including GPCR signaling and chromatin modification for RUNX1, and infection-related pathways and G alpha signaling for RUNX2 (**Supplemental Table 9**). We also examined the pathways enriched in the 1263 overlapping genes from the RUNX1 and RUNX2 two data sets and found that pathways involved in signal transduction, immune system, and axon guidance were enriched (**Figure 5D**). Examining the gene lists from ATAC-seq accessibility and gene set from CHEA Transcription Factor Targets databases for RUNX1, there were 16 genes that overlapped (*ABTB2, APBA2, ARHGEF7, ARPP21, ATG7, CNKSR3, CPA5, CTTNBP2, DISC1, FARS2, GABRA4, IPCEF1, NAB1, RELL1, SMARCA2, and TMEM39B*). Between the RUNX2 target gene set from the CHEA Transcription Factor Targets databases and genes located in differential accessible regions from ATAC-seq in fibroblasts, there were 9 overlapped (*ABTB2, CNKSR3, GABRA4, IPCEF1, NAB1, RANBP17, RELL1, SPATA16, and TRIM62*).

**Figure 5:**
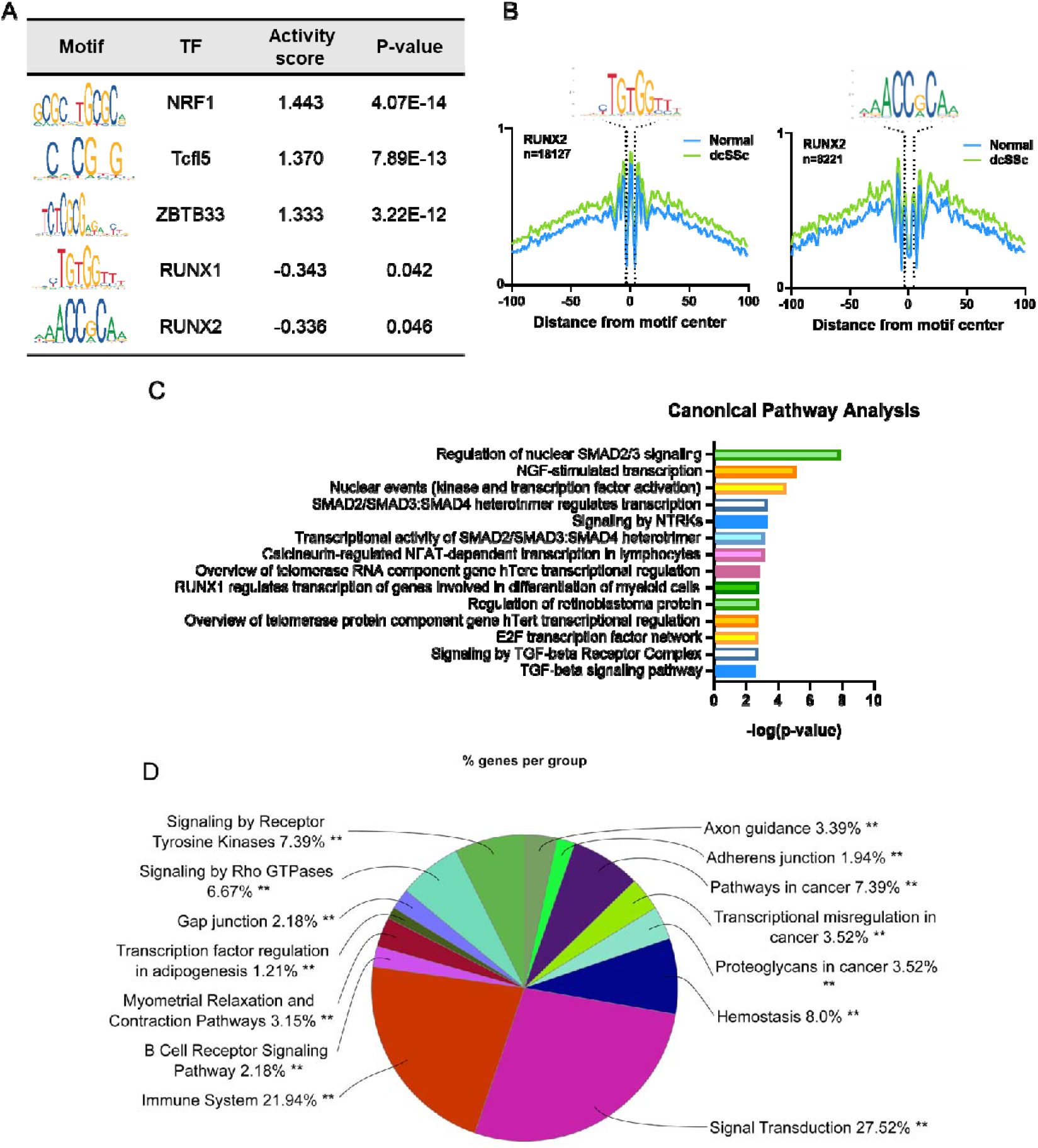
The most enriched transcription factor motifs and their target genes in dcSSc fibroblasts. (**A**) HINT-ATAC motif analysis of regions in normal and dcSSc fibroblasts. The top 3 transcription factors with significant differential activity values in fibroblasts are shown. The activity score indicates the difference in activity in normal compared to dcSSc fibroblasts (Normal-dcSSc). A positive score indicates increased transcription factor binding in normal fibroblasts, and a negative score indicates increased binding in dcSSc fibroblasts. (**B**) The footprint profiles of RUNX1 and RUNX2 generated from fibroblasts. (**C**) Pathway analysis of the putative transcription factors identified in fibroblasts. The top 14 are shown. (**D**) Pathway analysis of target genes regulated by both RUNX1 and RUNX2 identified using the CHEA Transcription Factor Targets database.

### Variable expression levels of transcription factors in dcSSc cells

To explore whether the putative transcription factors enriched in dcSSc ECs or fibroblasts were differentially expressed between normal and dcSSc cells, we performed qPCR to examine the mRNA levels of these transcription factors. In ECs, *ETV2* and *SNAI2* were significantly elevated in dcSSc ECs when compared to normal ECs (**Figure 6A**). We also examined *SNAI1*, which works alongside SNAI2 in many physiological pathways, in these cells. The expression of *SNAI1* was also significantly elevated in dcSSc ECs. In fibroblasts, the expression of *RUNX1* was not altered in dcSSc fibroblasts and normal fibroblasts while *RUNX2* was significantly elevated (**Figure 6B**).

**Figure 6:**
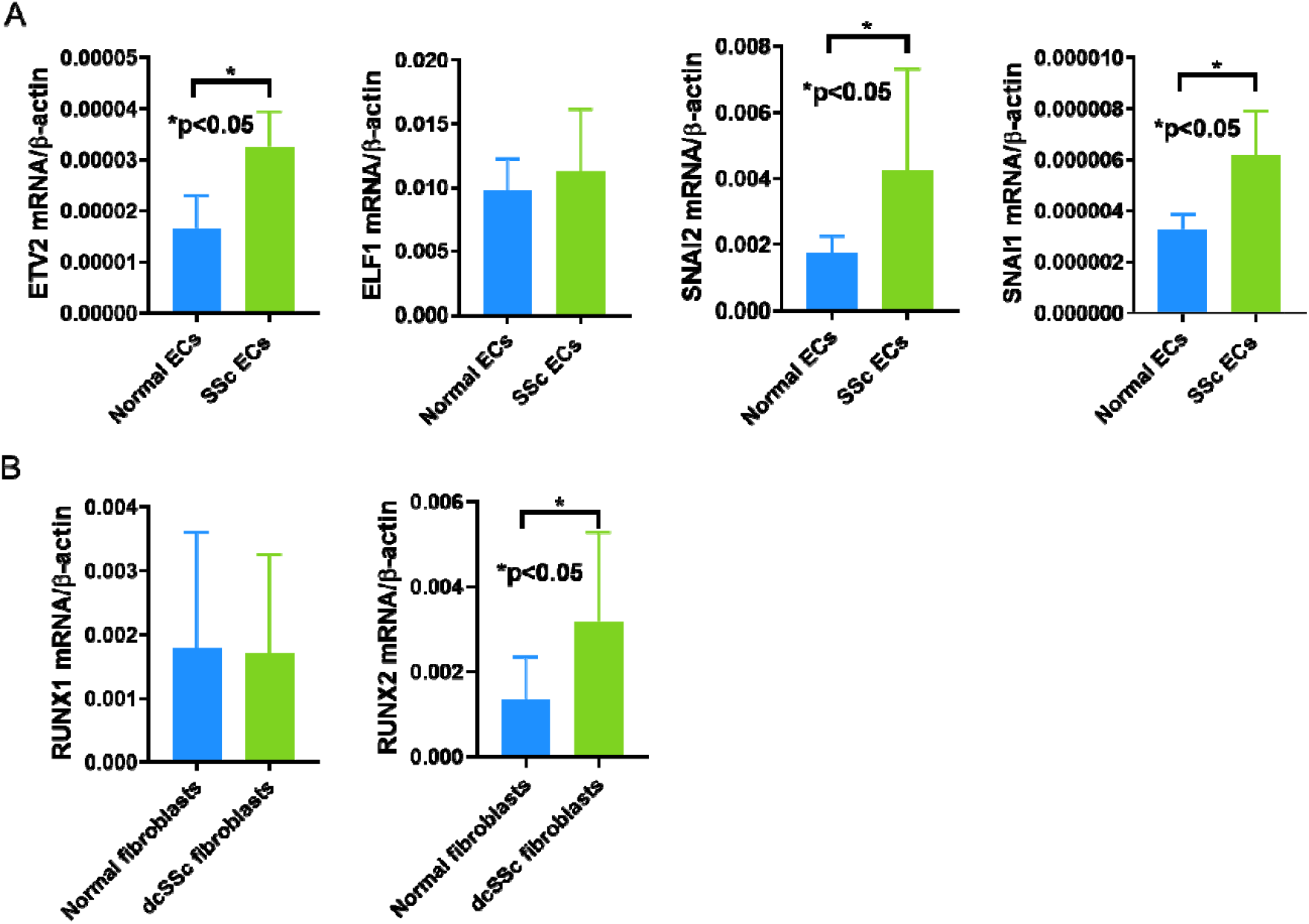
The mRNA expression levels of selected transcription factor genes in normal and dcSSc dermal (**A**) ECs (n=5) and (**B**) fibroblasts (n=12-16). Data are shown as mean +/-S.D.

## Discussion

It is known that environmental factors, which may alter epigenetic marks, contribute to the pathogenesis of SSc. In this work, we profiled changes of chromatin accessibility in ECs and fibroblasts isolated from dcSSc patients. We report a global reduction in chromatin accessibility in both cell types. In addition, we unveiled an unexpected neural signature in loci associated with differentially accessible regions from both cell types in dcSSc. By applying HINT-ATAC approaches, we defined for the first time the transcription factor footprints in dcSSc ECs and fibroblast at a genome-wide level. In all, this study uncovered genes and proteins that can serve as potential biomarkers or treatment targets for this disease.

Global changes in chromatin accessibility has been reported in many other diseases. In metastatic cancer cells genome-wide increase in chromatin accessibility has been reported to drive disease progression^24^. In contrast, neurodegenerative diseases such as age-related macular degeneration is associated with reduced chromatin accessibility^25^. Interestingly, aging has also been reported to be associated with global decrease in chromatin accessibility^26, 27^. Our results raise the possibility that the genome-wide reduction in chromatin accessibility could be a reflection of accelerated aging and senescence in dcSSc. Indeed, senescence is evident in SSc, as telomere shortening, which is a hallmark for senescence and aging has been reported^28, 29, 30^. Interestingly, the transcription factors in both cell types showed significant enrichment in pathways involved in hTERC and hTERT, which are members critical for telomerase regulation (**Figure 4C** and **5C**). In addition, the longevity-regulating pathway was enriched in overlapping genes between ones annotated from differentially accessible chromatin regions in dcSSc ECs and reported ETV2-target genes (**Figure 4D**).

The global decrease in chromatin accessibility in dcSSc ECs might be due to upregulation of histone deacetylases (HDACs). Histone lysine acetylation alters the charge on histone side chains resulting in an open chromatin configuration that allows transcription factors to bind and enhances gene transcription. We reported previously that in dcSSc ECs, several members of class II HDACs, including HDAC5, were significantly elevated ^6^. When we knocked down HDAC5 in dcSSc ECs, an increase in chromatin accessibility, as measured by ATAC-seq, was observed. This mechanism might also be relevant in dcSSc fibroblasts, as various HDACs, including HDAC1 and HDAC6, have been reported to be elevated in dcSSc fibroblasts when compared to their normal counterpart^31^.

Through pathway enrichment and gene ontology analysis of the genes annotated in differentially accessible chromatin regions, we observed an enrichment in genes that are involved in the nervous system. This was also one of the top enriched pathways in the transcription factors identified from HINT-ATAC in both ECs and fibroblasts (**Figure 4C** and **5C**). Although this neural signature might be surprising at first, the dysregulation of neuro-endothelial mechanisms has been suggested to be critical in initiating microvascular abnormalities such as Raynaud’s phenomenon in SSc^32^. Indeed, a disruptive peripheral nervous system, including morphological and functional changes, was reported in SSc ^33^. As the nervous and the vascular systems share anatomical patterns and many cellular mechanisms, it is not surprising that ECs express receptors for axon guidance molecules, which are grouped in 4 major families: Slit/Robo, semaphorin/plexin/neuropilin, Netrin/Unc5/DCC, and Ephrin/Eph^34^. It has been shown that in dcSSc ECs, dysregulation of these pathways led to defective angiogenesis^35, 36,37^. These results echo our pathway analysis where we detected significant enrichment in pathways involving axon guidance.

In addition to axon guidance pathways, we also observed enrichment in synaptic-related genes, including *CDH2, NLGN1*, and *NRXN*1. Besides their role in the nervous system, these genes are also critical in promoting EC angiogenesis. Loss- or gain-of-function experiments showed that CDH2 significantly promoted angiogenesis both *in vitro* and *in vivo*^38^. In a chicken chorioallantoic membrane system, a monoclonal recombinant antibody against NRXN1 blocked angiogenesis, whereas exogenous NLGN1 promoted angiogenesis^39^. We showed that the promoters of these genes are less accessible in dcSSc than normal ECs (**Figure 2B**). It is possible that these genes contribute to the dysregulated angiogenesis phenotype of dcSSc ECs.

Similar to what was observed in ECs, the enrichment of the genes associated with the nervous system in dcSSc fibroblasts also play relevant roles in fibrosis. *ERBB4*, which codes for receptor tyrosine-protein kinase ErbB-4, is a key cellular receptor for neuregulin growth factors. In mice, ErbB4 deletion accelerated renal fibrosis and renal injury^40^. In addition, the anti-fibrotic effect of neuregulin growth factor-1/ErbB4 pathway in the heart, skin, and lungs is linked to its anti-inflammatory activity in macrophages^41^.

Another example is *ENTPD1*, which codes for ectonucleoside triphosphate diphosphohydrolase 1 or CD39. In the nervous system, it hydrolyzes ATP to regulate neurotransmission. The deletion of ENTPD1/CD39 in mice promoted biliary injury and fibrosis by acting on gut-imprinted CD8+ T cells and macrophages^42, 43^. Since these two genes were annotated in differentially closing regions of the chromatin in dcSSc fibroblasts (**Figure 3B**), it is possible that they are downregulated in SSc and contribute to the exacerbated fibrosis phenotype in this disease.

When comparing the transcription factor footprints in dcSSc ECs, we found significant enrichment of *SNAI2* and *ETV2* in dcSSc ECs when compared to normal ECs. SNAI2 or Slug, is a neurogenic transcription factor that belongs to the Snail family of zinc finger transcription factors. It works with SNAI1 (or Snail) in processes involving epithelial- or endothelial-to-mesenchymal transition^44, 45^. In addition, it was found to be critical in angiogenic sprouting in ECs^46^. Here we report that both SNAI1 and SNAI2 were upregulated in dcSSc ECs. Since increased in endothelial-to-mesenchymal transition has been observed in dcSSc ECs^47^, the enrichment of SNAI2 might play a critical role in this process. ETV2, coding for transcription factor Ets variant 2, appears to be one of the key drivers for the enriched neuronal signal we observed in genes annotated in differentially accessible regions in dcSSc ECs. It is critical in promoting blood vessel formation as well as reprograming non-endothelial cells into cells displaying endothelial phenotypes^48^. As the regulatory network of ETV2 includes VEGF signaling, Notch pathways, MAPK signaling, and Ephrin signaling (**Supplemental Table 6**)^23^, its upregulation in dcSSc ECs would be expected to promote healthy EC function. The mechanism for the disconnect between upregulated ETV2 expression and impaired endothelial function in these cells warrants further examination.

The transcription factor RUNX2, which is enriched and upregulated in dcSSc dermal fibroblasts, belongs to the runt-related transcription factors. These RUNX proteins play critical roles in diverse cellular processes including proliferation, differentiation, epithelial–mesenchymal transition and inflammation. Studies have shown crosstalk between RUNX2 and pro-fibrotic signaling pathways, including TGFβ and Wnt pathways^49, 50^. Functionally, the effect of RUNX2 on fibrosis appears to be organ-specific. RUNX2-deficient mice showed exacerbated ureteral obstruction-induced kidney fibrosis, accompanied with enhanced TGFβ-signaling^51^. In contrast, RUNX2, which is significantly expressed in human type 2 diabetic aorta, induced aortic fibrosis and stiffening in mice^52^. In the lung, the involvement of RUNX2 in fibrogenesis is cell-type dependent^53^. Knockdown of RUNX2 in alveolar epithelial type II cells decreased profibrotic signals, whereas fibroblasts deficient in RUNX2 showed increased ECM production.

In this study, we provided novel information identifying open and closed chromatin regions in both ECs and fibroblasts in SSc. In addition, we characterized the transcription factor footprints in these cells and revealed the complex networks of transcription factors and their target genes, thereby allowing us to explore the potential mechanisms of dysregulated angiogenesis and enhanced fibrosis in this disease. While the decreased chromatin accessibility observed in ECs and fibroblasts might be a hallmark for dcSSc, it is not clear whether this observation contributes to the pathogenesis of the disease itself, or it is a mere representation of adaptive response for these cells to function in the disease environment. Further functional studies, accompanied with experiments identifying the molecular mechanisms that mediate the global changes in chromatin landscape in SSc will allow us to answer this question. This key information will identify new therapeutic targets for preventing or slowing disease progression for SSc.

## Data Availability

The data referred to in this manuscript are presented within the manuscript of the supplementary material provided.

